# Context-Aware Emergency Department Triage Using Pairwise Comparisons and Bradley-Terry Aggregation

**DOI:** 10.64898/2026.03.14.26348412

**Authors:** Philip Jarrett, Jonathan Reeder, Samuel McDonald, Deborah Diercks, Andrew Jamieson

## Abstract

**Objective:** To evaluate a ranking approach for emergency department (ED) waiting room prioritization that uses pairwise clinical comparisons aggregated via a Bradley-Terry model, and to assess its cross-site stability without site-specific training.

**Materials and Methods:** Using the Multimodal Clinical Monitoring in the Emergency Department (MC-MED) dataset (118,385 ED visits, Site A), we defined a composite deterioration outcome (intensive care unit [ICU] admission, intubation, vasopressor, ventilation, or death within 6 hours) and evaluated 7 queue-ordering policies across 1,000 simulated shifts. The primary endpoint was Recall@5 for deteriorators; secondary endpoints included area under the receiver operating characteristic curve (AUROC) and simulated time-to-provider (TTP) metrics. External validation used MIMIC-IV-ED (425,087 visits, Site B) with 500 shifts. Methods reported per TRIPOD-LLM.

**Results:** On MC-MED, BT-LLM-Enriched (Bradley-Terry ranking with a large language model [LLM] judge, GPT-4.1, using full diagnoses and medications) exceeded the Emergency Severity Index (ESI) on the primary endpoint: Recall@5 0.587 vs. 0.491 (p<0.001). XGBoost achieved Recall@5 0.648 but required large site-specific labeled training data. On external validation, supervised model performance attenuated (XGBoost AUROC 0.892 to 0.807) while BT-LLM-Enriched remained stable (0.826 to 0.831); the two were statistically indistinguishable on external data.

**Discussion:** Under external validation, supervised model performance attenuated while zero-shot LLM ranking remained stable, suggesting cross-site stability without requiring site-specific training data.

**Conclusion:** Pairwise ranking with an LLM judge significantly outperforms ESI-based ordering and remains stable across sites without local training, matching supervised models on external data.

## BACKGROUND AND SIGNIFICANCE

Emergency department (ED) triage systems primarily function as classifiers. The Emergency Severity Index (ESI)[1] assigns each patient to one of five acuity levels based on their individual presentation. The National Early Warning Score 2 (NEWS2)[2] and supervised machine learning (ML) risk models[3,4] similarly produce independent scores or categories for each patient in isolation. These tools inform the initial assessment of individual patient acuity, but they do not directly address a distinct operational question: among all patients currently in the waiting room, which one should be seen next?

Classification-based triage does not determine queue order. ESI assigns acuity levels but imposes no ordering within a level; among patients simultaneously classified as ESI-3, the default ordering is typically first-in-first-out (FIFO).[1] Post hoc sorting of independent risk scores produces an ordering but does not incorporate information about other patients in the queue. Supervised risk models additionally require large site-specific labeled training sets that may not be available at all institutions, and their generalizability across sites with different patient populations remains uncertain.[3,4] Systematic reviews of triage performance have identified substantial room for improvement in identifying patients at risk for adverse outcomes, with sensitivity for critical illness ranging from 36% to 91% depending on the condition and triage system.[7] Given that ED delays are independently associated with increased mortality[5,6], optimizing the order in which patients are seen has direct safety implications.

Determining which patient should be seen next is fundamentally a ranking problem. Rather than assigning each patient an independent score, a ranking approach determines each patient’s position relative to the other patients currently waiting. Pairwise comparisons offer a natural mechanism for this: each new arrival is compared against selected patients already in the queue, and the outcomes are aggregated to produce an ordering that encodes queue context directly. Although absolute risk models can be used to sort patients by predicted probability of deterioration, such models evaluate each patient independently and require careful calibration across institutions. Queue prioritization, however, is inherently a relative decision among patients simultaneously competing for clinical attention. A ranking formulation allows prioritization to be determined directly from pairwise comparisons and aligns the modeling objective with the operational decision.

Pairwise comparisons can be aggregated using established ranking models such as the Bradley-Terry model[9]. This model estimates a latent severity score for each patient from pairwise preferences, producing a consistent ranking even when individual comparisons are non-transitive[8]. Bradley-Terry aggregation has been applied at scale for large language model (LLM) evaluation[10,11] and has recently been proposed for prioritizing patient portal messages by medical urgency[12], but has not previously been applied to ED queue prioritization. Pairwise comparison also enables the use of large language models as comparison functions, allowing prioritization decisions to incorporate unstructured clinical context without requiring probability calibration.[13,14,15] The comparison function is modular: it may be a vital-sign heuristic, a trained classifier, or an LLM that can reason over the full clinical context of a patient’s triage presentation.

We evaluated this ranking approach for ED waiting room prioritization across two institutions, testing pairwise comparison functions ranging from a deterministic vital-sign heuristic to GPT-4.1 with detailed diagnosis and medication information.

## OBJECTIVE

The primary objective was to evaluate whether reframing ED queue prioritization as a ranking task, implemented via pairwise comparison and Bradley-Terry aggregation, improves top-of-queue identification of patients at risk of near-term deterioration compared with classification-based ordering (ESI, NEWS2, supervised ML). The secondary objectives were to determine how the richness of clinical information available to the pairwise judge affects ranking quality, and to assess the zero-shot cross-site stability of the framework through external validation.

## MATERIALS AND METHODS

### Development dataset

We used the Multimodal Clinical Monitoring in the Emergency Department (MC-MED) dataset[16,17], a de-identified, event-level dataset from a single academic medical center (Site A) comprising 118,385 adult ED visits. The dataset includes visit-level demographics, triage vital signs, structured chief complaint codes, and ESI acuity assignments; timestamped orders (3.3 million records); laboratory results (5.7 million); continuous vital sign measurements (47.8 million); home medication records with drug names and therapeutic classes; and past medical history (PMH) coded in ICD-10 with diagnosis descriptions. Analyses used a chronological test split (12,020 visits) with model training on the chronological training split (81,588 visits).

### External validation dataset

For external validation, we used the MIMIC-IV-ED dataset[18] linked to MIMIC-IV[19,20] (Site B), comprising 425,087 adult ED visits from a separate academic medical center. Triage data (vital signs, ESI acuity, free-text chief complaint, pain score) were obtained from the MIMIC-IV-ED triage table. PMH was reconstructed from billed diagnoses of prior hospital admissions for the same patient, linked via the MIMIC-IV diagnoses_icd table. Active medications were obtained from the MIMIC-IV-ED medication reconciliation table. Outcomes were derived from MIMIC-IV hospital and intensive care unit (ICU) modules using the same composite definition as the development dataset, linked by hospital admission identifier.

Both datasets are available through PhysioNet[21] and were accessed under the Credentialed Health Data License (version 1.5.0), which requires completion of recognized training in human research subject protection and Health Insurance Portability and Accountability Act (HIPAA) regulations, as well as execution of a data use agreement. Because the data are de-identified in accordance with the HIPAA Safe Harbor standard, this study did not constitute human subjects research and was not subject to institutional review board oversight.

### Triage capsule construction

For each visit, we constructed a triage capsule limited to information available at or before triage. The capsule contained: age and sex; arrival mode; ESI acuity level; triage vital signs (heart rate, blood pressure, respiratory rate, oxygen saturation, temperature, pain score); chief complaint; and binary high-risk flags for hypotension (systolic blood pressure <90 mmHg), hypoxia (SpO2 <92%), tachypnea, tachycardia, fever, hypothermia, altered mental status (chief complaint proxy), chest pain, and sepsis concern (two or more systemic inflammatory response syndrome [SIRS] criteria). Missingness indicators were included for each vital.

The capsule was serialized in two text formats for LLM pairwise comparisons. The structured format (∼200 tokens per patient) included vital signs, chief complaint, a count of PMH entries, and four binary home medication flags (anticoagulant, insulin, immunosuppressant, antineoplastic). The enriched format replaced these summary fields with actual diagnosis descriptions and active medication names, providing clinical specificity beyond what binary flags and counts can convey (∼350 tokens per patient). A 32-element numeric feature vector was used for the ML baseline.

### Outcome definition

The primary outcome was a composite indicator of clinical deterioration defined as any of the following within 6 hours of ED arrival — ICU admission, intubation order, vasopressor initiation, or mechanical ventilation order — or in-hospital death.[22,23] The composite deterioration rate was 2.5% in the MC-MED test cohort (301 of 12,020 visits) and 4.6% in MIMIC-IV-ED (19,507 of 425,087 visits); the contribution of each component is detailed in Supplementary Table 1.

### Waiting room simulation

Per-patient date shifting in both datasets precludes reconstruction of concurrent waiting room states, necessitating simulated shift scenarios. We generated shifts by sampling from each test cohort without replacement. Time-to-provider (TTP) was modeled as proportional to queue position: for a shift of n patients, the patient at queue position k was assigned a simulated wait of (k/n) × 480 minutes. Because TTP is a monotonic function of queue position, relative differences between policies reflect differences in ranking order regardless of the specific throughput function; a sensitivity analysis confirmed that the relative ordering of all policies was preserved under convex and concave wait-time functions (Supplementary Figure 1). Because real waiting-room progression could not be reconstructed from the available datasets, simulated TTP is presented as a secondary operational illustration rather than a direct measure of real waiting times. For MC-MED, we generated 1,000 shifts stratified by census level (333 low-census at 25 patients, 333 medium at 40, 334 high at 60), yielding 41,685 patient-encounters per policy. For MIMIC-IV-ED, we generated 500 shifts with identical stratification (166/166/168), yielding 20,820 patient-encounters per policy. Patient characteristics in simulated shifts were comparable to the full test cohorts (Supplementary Table 2).

### Queue-ordering policies

We evaluated 7 policies representing progressively richer comparison capabilities (Supplementary Table 3; Figure 1). Standard-of-care baselines: (1) FIFO, (2) ESI, (3) NEWS2.[2] Supervised ML comparator: (4) XGBoost, trained on 81,588 MC-MED encounters using 32 capsule features[24] and applied without retraining to MIMIC-IV-ED. Within the Bradley-Terry pairwise framework, we evaluated three progressively richer comparison functions: (5) BT-Heuristic, using a deterministic NEWS2-based judge; (6) BT-LLM, using GPT-4.1 with the structured capsule; and (7) BT-LLM-Enriched, using GPT-4.1 with the enriched capsule containing full diagnosis descriptions and medication names. GPT-4.1 (version gpt-4.1-2025-04-14, developed by OpenAI) was accessed securely through Microsoft Azure.

**Figure 1.**
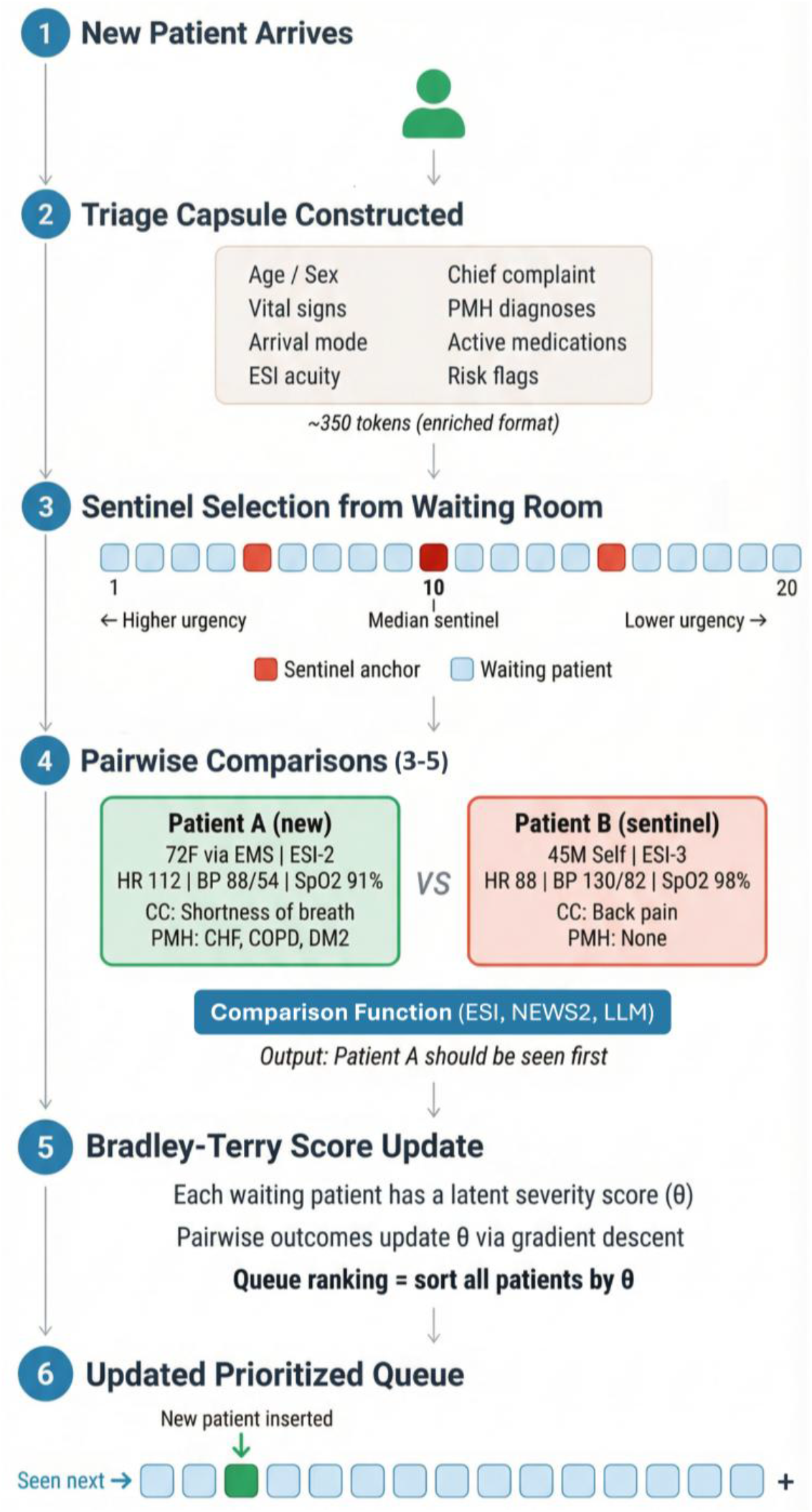
Algorithm schematic showing the pairwise ranking pipeline. When a new patient arrives (step 1), a triage capsule is constructed from available clinical data (step 2). Sentinel patients are selected from the current waiting room at quantiles of the severity distribution (step 3). The new patient is compared pairwise against each sentinel by a pluggable comparison function (step 4), and comparison outcomes update latent severity scores (θ) via the Bradley-Terry model (step 5). The queue is re-sorted by severity score to produce an updated prioritized order (step 6). Abbreviations: BP, blood pressure; BT, Bradley-Terry; CC, chief complaint; CHF, congestive heart failure; COPD, chronic obstructive pulmonary disease; DM2, type 2 diabetes mellitus; EMS, emergency medical services; ESI, Emergency Severity Index; HR, heart rate; LLM, large language model; NEWS2, National Early Warning Score 2; PMH, past medical history; SpO2, peripheral oxygen saturation; θ, latent severity score. Alt text: Flowchart showing the pipeline from patient arrival through triage capsule construction, sentinel selection, pairwise comparison with a pluggable judge, Bradley-Terry model update, severity ranking, and prioritized queue output.

Pairwise comparison outcomes were aggregated using the Bradley-Terry model[9], which estimates latent severity scores from pairwise preferences to produce a consistent ranking. Update equations, confidence weighting, convergence properties, and hyperparameter sensitivity analyses are detailed in Supplementary Appendix A. New patients were compared against 3–5 sentinel anchors selected at quantiles of the current severity distribution, with 2 additional comparisons for patients entering the top 5 (top-k reinforcement), yielding 3–7 total comparisons per arrival.

For each pair, the LLM was prompted to return a structured JavaScript Object Notation (JSON) object specifying which patient should be seen first (full prompt template in Supplementary Material). Temperature was set to 0, and no free-text explanations were elicited or stored. External validation on MIMIC-IV-ED included all standard-of-care baselines, XGBoost, BT-Heuristic, and BT-LLM-Enriched. BT-LLM was not separately evaluated on external data because BT-LLM-Enriched significantly outperformed it on the development dataset, establishing the enriched capsule as the top-performing configuration within the ranking framework; external validation was designed to test the cross-site stability of this best configuration against classification-based ordering and supervised ML.

### Primary endpoint and statistical analysis

Methods and results are reported in accordance with the TRIPOD-LLM reporting guideline.[25] The primary endpoint was Recall@5 for deteriorators, defined as the fraction of deteriorators ranked in positions 1–5 per shift, for the comparison of BT-LLM-Enriched versus ESI. Because queue prioritization is inherently a top-of-ranking problem, Recall@5 directly evaluates whether the framework places the highest-risk patients at the front of the queue, aligning the endpoint with the operational decision.[7] Secondary endpoints included area under the receiver operating characteristic curve (AUROC), which evaluates global discrimination across the full risk spectrum, and simulated TTP metrics (median and 95th-percentile TTP for deteriorators), which provide an operational illustration of the ranking’s downstream impact on wait times under an assumed throughput model. All comparisons use paired t-tests on shift-level metrics with 95% bootstrap confidence intervals (500 resamples). Shifts without deterioration events were excluded.

## RESULTS

### Development dataset (MC-MED)

Table 1 presents the MC-MED results. BT-LLM-Enriched significantly exceeded ESI on the primary endpoint: Recall@5 was 9.6 percentage points higher (0.587 vs. 0.491; paired difference 95% CI: +6.1 to +13.2; p<0.001; Figure 2). On secondary endpoints, AUROC was 0.826 for BT-LLM-Enriched versus 0.811 for ESI (p=0.09; Figure 3), consistent with the expectation that AUROC, which evaluates global discrimination, may be less sensitive to improvements concentrated at the top of the ranking. Secondary TTP metrics also favored BT-LLM-Enriched: median TTP for deteriorators was 9.3 minutes shorter (79 vs. 88 minutes; paired difference 95% CI: −17.5 to −1.8; p=0.026) and the 95th-percentile tail wait was 116 minutes versus 122 minutes for ESI (Supplementary Figure 2).

**Table 1.**
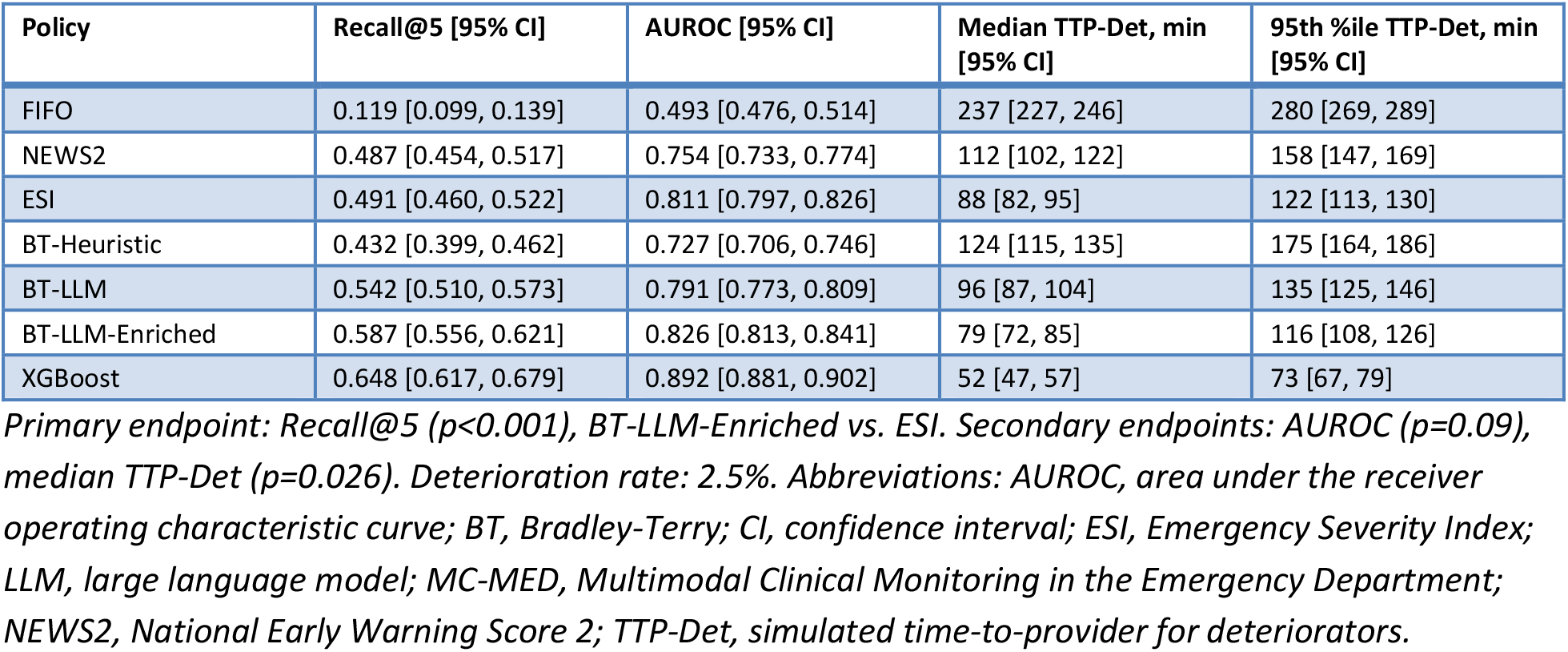
Development dataset results: MC-MED (1,000 simulated shifts, Site A).

**Figure 2.**
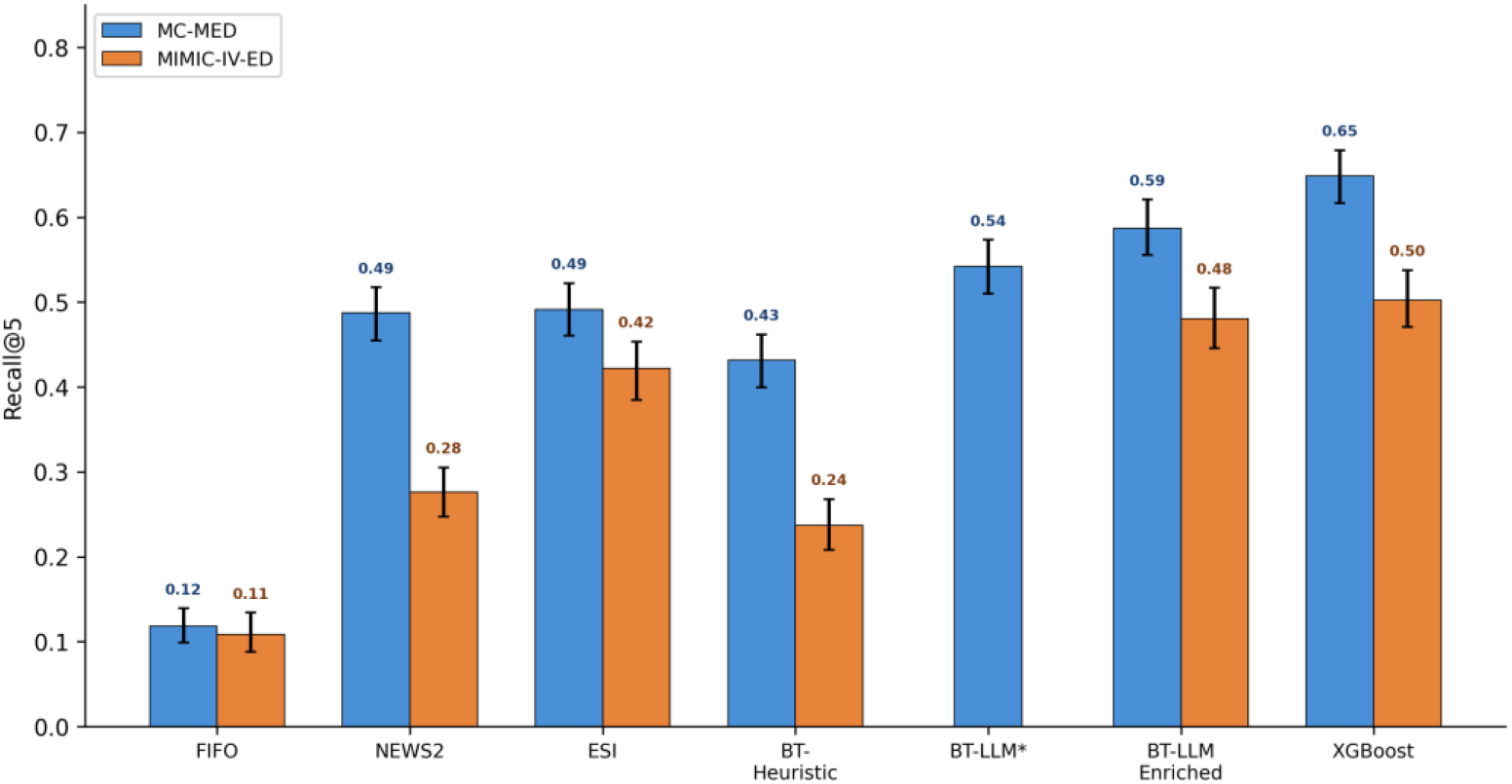
Recall@5 for deteriorators across development (MC-MED, blue) and external validation (MIMIC-IV-ED, orange) datasets (primary endpoint). Recall@5 is the fraction of deteriorators ranked in positions 1–5 per shift. Error bars show 95% bootstrap confidence intervals. Asterisk (*) indicates BT-LLM was not evaluated on external validation; only BT-LLM-Enriched was used. Abbreviations: BT, Bradley-Terry; ESI, Emergency Severity Index; FIFO, first-in-first-out; LLM, large language model; MC-MED, Multimodal Clinical Monitoring in the Emergency Department; MIMIC-IV-ED, Medical Information Mart for Intensive Care IV Emergency Department; NEWS2, National Early Warning Score 2. Alt text: Grouped bar chart of Recall at 5 for 7 policies across MC-MED and MIMIC-IV-ED. BT-LLM-Enriched achieves 0.587 on MC-MED and 0.481 on MIMIC-IV-ED, exceeding ESI at both sites. XGBoost is highest on MC-MED at 0.648.

**Figure 3.**
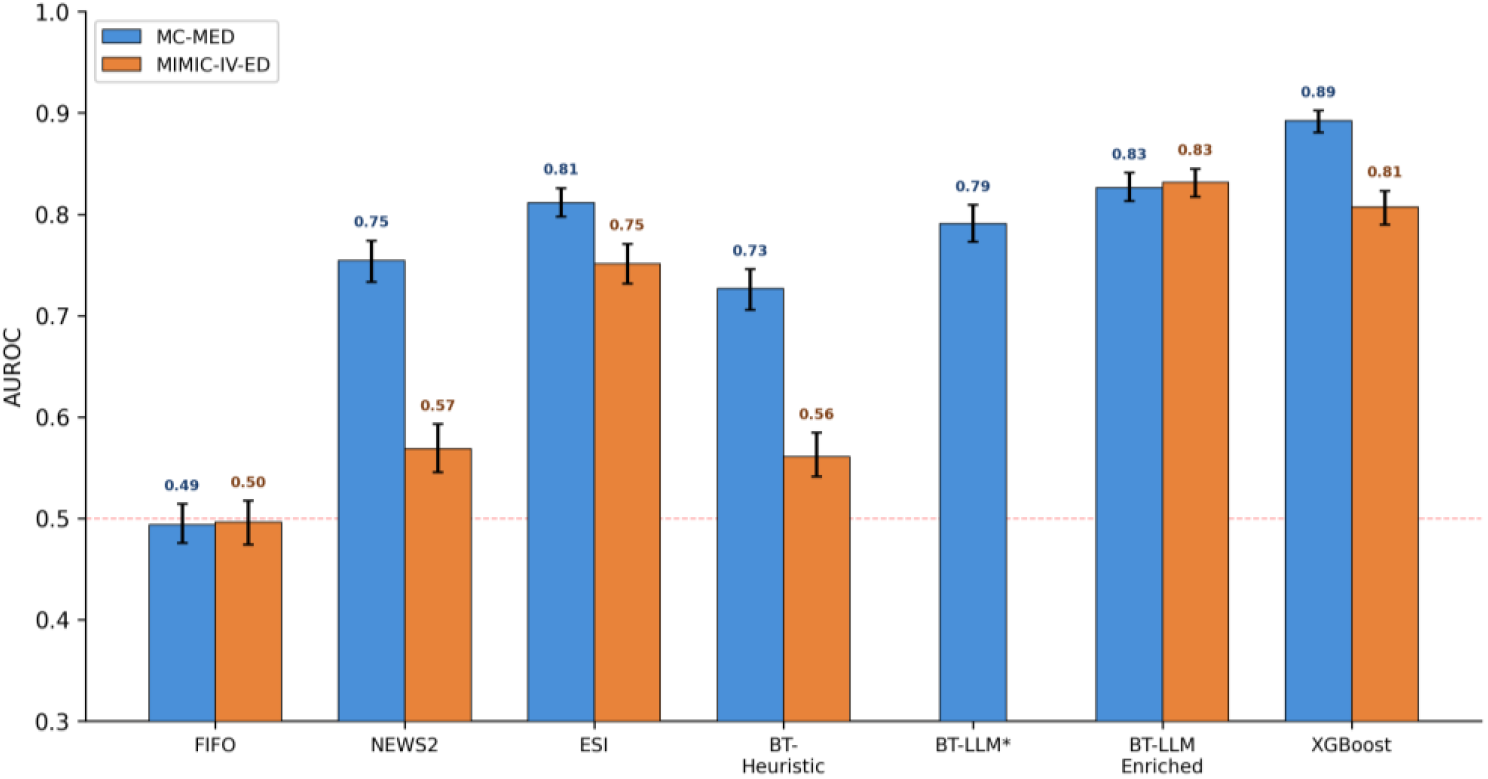
AUROC for deterioration prediction across development (MC-MED, blue) and external validation (MIMIC-IV-ED, orange) datasets (secondary endpoint). Dashed red line indicates chance discrimination (AUROC = 0.5). Error bars show 95% bootstrap confidence intervals. Asterisk (*) indicates BT-LLM was not evaluated on external validation; only BT-LLM-Enriched was used. Abbreviations: AUROC, area under the receiver operating characteristic curve; BT, Bradley-Terry; ESI, Emergency Severity Index; FIFO, first-in-first-out; LLM, large language model; MC-MED, Multimodal Clinical Monitoring in the Emergency Department; MIMIC-IV-ED, Medical Information Mart for Intensive Care IV Emergency Department; NEWS2, National Early Warning Score 2. Alt text: Grouped bar chart of AUROC for 7 policies across MC-MED and MIMIC-IV-ED. XGBoost is highest on MC-MED at 0.892 but attenuates to 0.807 on MIMIC-IV-ED. BT-LLM-Enriched achieves 0.826 and 0.831 across sites.

The supervised XGBoost comparator, trained on 81,588 encounters from the same institution, achieved the highest Recall@5 (0.648), lowest median TTP (52 minutes), and lowest tail wait (73 minutes).

Within the Bradley-Terry framework, each increase in comparison function capability produced significant gains: BT-Heuristic Recall@5 0.432, BT-LLM 0.542 (paired difference +11.0 [+7.7, +14.2]; p<0.001), BT-LLM-Enriched 0.587 (paired difference +4.5 [+1.9, +7.2]; p=0.001 vs. BT-LLM).

### External validation (MIMIC-IV-ED)

Table 2 presents the MIMIC-IV-ED results. BT-LLM-Enriched exceeded ESI on Recall@5 (0.481 vs. 0.422; paired difference +3.8 [−1.1, +9.0]; p=0.15), though this did not reach significance on the external dataset. On secondary endpoints, BT-LLM-Enriched significantly exceeded ESI on AUROC (0.831 vs. 0.751; paired difference +7.5 [+4.9, +10.0]; p<0.001) and simulated TTP metrics (median TTP 77 vs. 112 minutes; paired difference −31.2 [−43.2, −19.2]; p<0.001; 95th-percentile tail wait 129 vs. 184 minutes; paired difference −54.7 [−71.6, −37.5]; p<0.001).

**Table 2.** External validation results: MIMIC-IV-ED (500 simulated shifts, Site B).

| Policy | Recall@5 [95% CI] | AUROC [95% CI] | Median TTP-Det, min [95% CI] | 95th %ile TTP-Det, min [95% CI] |
| --- | --- | --- | --- | --- |
| FIFO | 0.109 [0.087, 0.131] | 0.496 [0.476, 0.516] | 235 [225, 246] | 304 [294, 316] |
| NEWS2 | 0.276 [0.248, 0.307] | 0.569 [0.544, 0.593] | 202 [191, 215] | 286 [273, 299] |
| ESI | 0.422 [0.388, 0.457] | 0.751 [0.733, 0.770] | 112 [103, 121] | 184 [172, 195] |
| BT-Heuristic | 0.237 [0.206, 0.266] | 0.561 [0.538, 0.584] | 204 [192, 216] | 288 [276, 302] |
| BT-LLM-Enriched | 0.481 [0.444, 0.516] | 0.831 [0.818, 0.844] | 77 [71, 83] | 129 [120, 138] |
| XGBoost (Site A trained) | 0.502 [0.468, 0.537] | 0.807 [0.790, 0.823] | 86 [78, 94] | 154 [143, 165] |

*Primary endpoint: Recall@5 ( $p=0.15$ ), BT-LLM-Enriched vs. ESI. Secondary endpoints: AUROC ( $p<0.001$ ), median TTP-Det ( $p<0.001$ ). BT-LLM-Enriched vs. XGBoost: all metrics $p>0.05$ except 95th %ile TTP ( $p=0.005$ ). Deterioration rate: 4.6%. XGBoost applied without retraining from Site A model. BT-LLM and BT-LLM-Enriched used same prompt and model (GPT-4.1) as development dataset. BT-LLM not separately evaluated on MIMIC-IV-ED. Abbreviations: AUROC, area under the receiver operating characteristic curve; BT, Bradley-Terry; CI, confidence interval; ESI, Emergency Severity Index; LLM, large language model; MIMIC-IV-ED, Medical Information Mart for Intensive Care IV Emergency Department; NEWS2, National Early Warning Score 2; TTP-Det, simulated time-to-provider for deteriorators.*

Under external validation, supervised model performance attenuated while LLM performance remained stable (Figure 4). XGBoost, applied without retraining, achieved AUROC 0.807 (vs. 0.892 on MC-MED) and Recall@5 0.502 (vs. 0.648). BT-LLM-Enriched achieved AUROC 0.831 (vs. 0.826 on MC-MED) and Recall@5 0.481 (vs. 0.587). On the external dataset, BT-LLM-Enriched and XGBoost were statistically indistinguishable on all metrics (Recall@5 paired difference −0.1 [−6.9, +7.2], p=0.97; AUROC paired difference +2.3 [−0.1, +4.9], p=0.07; TTP paired difference −7.3 [−19.0, +3.9], p=0.22). BT-LLM-Enriched achieved significantly lower 95th-percentile tail wait than XGBoost (129 vs. 154 minutes; paired difference −24.5 [−42.0, −7.2]; p=0.005).

**Figure 4.**
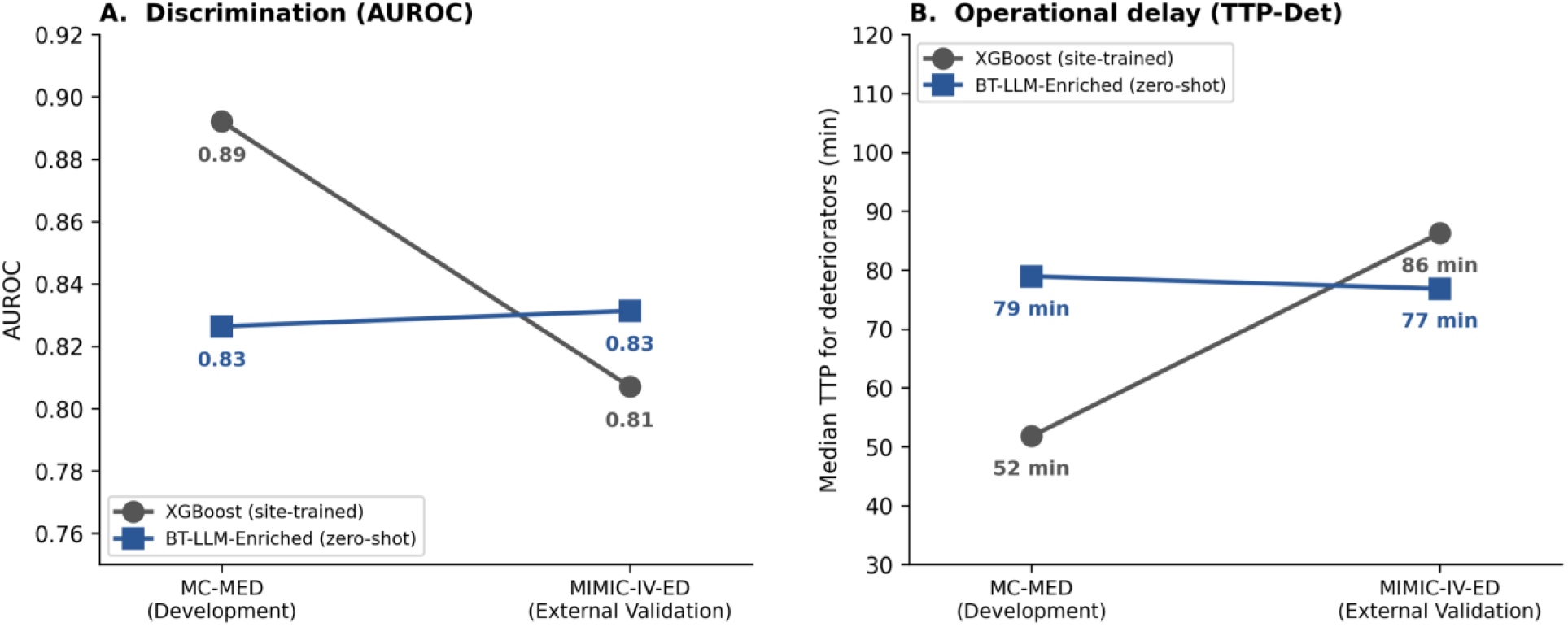
Cross-site comparison: AUROC and median TTP-Det for BT-LLM-Enriched and XGBoost across development (MC-MED) and external validation (MIMIC-IV-ED) datasets. XGBoost (applied without retraining) performance attenuates on external data while zero-shot BT-LLM-Enriched remains stable. Abbreviations: AUROC, area under the receiver operating characteristic curve; BT, Bradley-Terry; LLM, large language model; MC-MED, Multimodal Clinical Monitoring in the Emergency Department; MIMIC-IV-ED, Medical Information Mart for Intensive Care IV Emergency Department; TTP-Det, simulated time-to-provider for deteriorators. Alt text: Paired comparison showing XGBoost AUROC declining from 0.892 to 0.807 across sites while BT-LLM-Enriched remains stable at 0.826 and 0.831.

### Subgroup analysis (MIMIC-IV-ED)

Per-patient ranking data from the external validation enabled subgroup-stratified comparison of top-5 identification rates across policies. ESI-based ordering showed a significant disparity by arrival mode: self-arriving deteriorators were placed in the top 5 at a rate of 48.2%, compared with 36.4% for emergency medical services (EMS) arrivals (chi-squared p=0.0004). Under BT-LLM-Enriched, this disparity was not observed (EMS 48.6%, self-arrival 44.0%; p=0.14). No significant disparity by age group was observed under either ESI (p=0.83) or BT-LLM-Enriched (p=0.18). These findings are consistent with prior literature documenting undertriage patterns associated with arrival mode[26] and age.[27,28] Development-site subgroup identification rates and deterioration base rates are provided in Supplementary Tables 4 and 5, respectively. Racial subgroup cell sizes were insufficient for statistical comparison, though directionally, Black/African American deteriorators had a higher top-5 rate under BT-LLM-Enriched (50.8%) than under ESI (37.8%).

### Computational cost

Each patient arrival required 3–5 pairwise LLM comparisons under the sentinel strategy. At current GPT-4.1 pricing ($2/million input tokens, $8/million output tokens), this corresponds to approximately $0.01 per patient arrival. For an ED processing 150 patients per day, the estimated daily LLM inference cost would be approximately $1.50, or roughly $550 per year. These estimates reflect pricing at time of study; model pricing is subject to change, though the trend has been downward. BT-Heuristic incurs zero inference cost and may be suitable where LLM access is infeasible.

## DISCUSSION

This study introduces and externally validates a pairwise ranking approach for ED waiting room prioritization. Four principal findings emerged.

First, BT-LLM-Enriched significantly outperformed ESI on the primary endpoint, Recall@5, at the development site (p<0.001) and showed consistent improvement at the external site (Tables 1 and 2). The ranking approach concentrates its advantage at the top of the queue, where operational impact is greatest, consistent with the expected behavior of a pairwise aggregation approach designed for top-of-queue identification.

Second, the richness of clinical information provided to the pairwise judge was the most impactful design decision. Each step from vital-sign heuristic to LLM with structured capsules to LLM with enriched capsules produced significant gains (Table 1). The enriched triage summary enabled the LLM to reason about specific clinical patterns, such as the combination of a benign chief complaint with a high-risk past medical history, that binary summary flags cannot convey.

Third, under external validation, supervised model performance attenuated while zero-shot LLM ranking remained stable (Table 2). XGBoost AUROC declined substantially across sites, whereas BT-LLM-Enriched was essentially unchanged; on the external dataset, the two approaches were statistically indistinguishable. Several factors may contribute to this pattern, including distribution shift, differences in outcome prevalence between sites, and site-specific training-data dependence in the supervised model. The stability of LLM-based performance is consistent with zero-shot approaches leveraging broader clinical priors that are less dependent on local training distributions,[29] though the relative contribution of each factor cannot be isolated in this study design. Supervised models with site-specific adaptation (e.g., Platt scaling or limited retraining) may narrow this gap; such recalibration experiments were outside the scope of this study, which focused on zero-shot cross-site stability rather than optimized supervised transfer. BT-LLM-Enriched also achieved significantly lower worst-case tail wait than XGBoost on external validation (p=0.005), suggesting a potential safety advantage in the tail of the distribution.

Fourth, BT-LLM-Enriched significantly outperformed ESI despite operating on structured electronic health record (EHR) data rather than direct patient contact. Triage nurses face cognitive demands including decision fatigue[30] and susceptibility to anchoring biases.[31] This approach is designed as a decision-support layer that augments, rather than replaces, nurse triage: a preliminary severity ranking from EHR data at registration could reduce cognitive burden while preserving clinical override.[32,33] This hypothesis requires prospective evaluation of provider interaction with ranked queues. On the external validation dataset, ESI-based ordering demonstrated a significant disparity by arrival mode, with EMS-arriving deteriorators identified at a lower rate than self-arrivals. This disparity was not observed under BT-LLM-Enriched, consistent with the framework’s reliance on clinical data rather than contextual cues such as transport mode that may introduce bias into human triage decisions. Prior work has documented racial and ethnic disparities in ESI assignment[34] and within-acuity prioritization[35]; racial subgroup cell sizes in this study were insufficient for definitive analysis, and prospective evaluation across diverse populations remains an important next step.

Because simulated TTP is a deterministic transform of queue position under an assumed throughput model, TTP results should be interpreted as illustrative of the ranking’s operational impact rather than precise estimates of ED waiting times. The consistent direction of TTP differences across sites corroborates the primary ranking-based findings, but absolute TTP values are contingent on the assumed throughput function and should not be interpreted as operational predictions.

More broadly, the distinction between classification and ranking as decision support paradigms extends beyond ED triage. Many clinical operational decisions, including ICU prioritization during surge capacity, transplant allocation, and radiology worklist ordering, require relative prioritization among competing candidates. This pairwise approach is not specific to the ED and may be useful wherever clinicians must prioritize competing patients for limited resources.

### Limitations

The synthetic waiting room simulation does not capture real temporal dynamics such as time-varying crowding or provider availability. The ranking architecture itself does not depend on simulation and could be evaluated prospectively; the simulation was required only to estimate operational metrics under counterfactual queue orderings. Real rooming decisions were not observed in either dataset; provider availability and clinician judgment, which may influence prioritization independent of acuity, are not modeled. ESI-based ordering may not strictly determine rooming order in practice, as clinician judgment and bed availability can influence which patient is taken next. Simulated TTP values reflect queue position under an assumed proportional throughput model; absolute wait times should be interpreted as relative comparisons between policies rather than operational predictions. The composite outcome aggregates heterogeneous clinical events (Supplementary Table 1). PMH sources differed between datasets (dedicated problem list in MC-MED; prior admission diagnoses in MIMIC-IV), and triage vital missingness was higher in MIMIC-IV-ED (4–5% vs. <1%). The LLM’s judgments may encode biases from pretraining data, and although the subgroup analysis on MIMIC-IV-ED did not reveal significant disparities under BT-LLM-Enriched, cell sizes for racial subgroups were small and prospective evaluation across diverse populations remains necessary. This study evaluated a fixed set of policies; adaptive approaches that learn from outcomes over time were not tested. This work represents a methodological proof-of-concept; no human-in-the-loop evaluation, workflow integration modeling, or provider acceptability assessment was performed, and these would be required before clinical deployment.

## CONCLUSION

Reframing ED queue prioritization as a ranking task, using pairwise comparisons aggregated by a Bradley-Terry model, improves identification of deteriorating patients compared with ordering based on independent risk scores. Using an LLM with enriched clinical capsules, this approach significantly outperformed ESI at both institutions and matched a site-trained supervised model on external validation without requiring local training data. Under external validation, supervised model performance attenuated while the LLM-based framework remained stable, demonstrating zero-shot cross-site stability. Supervised models with site-specific adaptation (e.g., recalibration or limited retraining) may narrow this gap, but the zero-shot approach eliminates the requirement for local labeled data entirely. This approach offers a modular method for clinical queue prioritization that warrants prospective evaluation.

## Data Availability

All results created during this research are derivatives of Physionet datasets, which the authors of this work are unable to share. Code that demonstrates the pairwise comparison using Bradley-Terry aggregation may be provided on an individual basis upon reasonable request by emailing the corresponding author.

## ACKNOWLEDGMENTS

During the preparation of this work, the authors used Claude Opus 4.6 to execute the planned data visualizations in Python and draft portions of this article. After using this tool, the authors reviewed and edited the content as needed and take full responsibility for the content of the published article.

## DATA AND CODE AVAILABILITY

Both datasets are publicly available through PhysioNet (MC-MED, https://doi.org/10.13026/jz99-4j81; MIMIC-IV-ED, https://doi.org/10.13026/5ntk-km72) under the Credentialed Health Data License. Code for the Bradley-Terry aggregation and pairwise comparison framework is available from the corresponding author upon reasonable request. Raw LLM outputs cannot be shared because they constitute derived data from the PhysioNet datasets and are subject to the original data use agreements.

