## Supplements for "Context-Aware Emergency Department Triage Using Pairwise Comparisons and Bradley-Terry Aggregation"

**Supplementary Table 1. Composite deterioration outcome by component.**

| Component | Within 6 hours | Any time | % of composite (N=301) |
|---|---|---|---|
| ICU admission | 220 (1.83%) | 251 (2.09%) | 73.1% |
| In-hospital death | 104 (0.87%) | 104 (0.87%) | 34.6% |
| Vasopressor initiation | 53 (0.44%) | 59 (0.49%) | 17.6% |
| Mechanical ventilation | 50 (0.42%) | 51 (0.42%) | 16.6% |
| Intubation | 44 (0.37%) | 46 (0.38%) | 14.6% |
| **Composite** | **301 (2.50%)** | | |

*Percentages in the "% of composite" column exceed 100% because patients may qualify by multiple criteria. Among the 301 composite-positive patients, 206 (68%) qualified by exactly 1 criterion, 42 (14%) by 2, 34 (11%) by 3, and 19 (6%) by 4 or more. Abbreviation: ICU, intensive care unit.*

**Supplementary Table 2. Simulated shift characteristics compared with test cohort.**

| Characteristic | Full test cohort (N=12,020) | Sampled into shifts (N=11,630) |
|---|---|---|
| Age, mean (SD) | 53.0 (20.5) | 53.0 (20.5) |
| Female, % | 53.3 | 53.4 |
| EMS arrival, % | 19.4 | 19.4 |
| Deterioration rate, % | 2.50 | 2.48 |
| ESI 1, % | 0.8 | 0.8 |
| ESI 2, % | 26.5 | 26.6 |
| ESI 3, % | 62.4 | 62.3 |
| ESI 4, % | 9.8 | 9.8 |
| ESI 5, % | 0.5 | 0.5 |
| Heart rate, mean (SD) | 86.2 (18.5) | 86.2 (18.4) |
| Systolic BP, mean (SD), mmHg | 135.6 (22.1) | 135.7 (22.1) |
| Respiratory rate, mean (SD) | 18.0 (2.6) | 18.0 (2.5) |
| $SpO_2$, mean (SD), % | 98.2 (2.1) | 98.2 (2.1) |
| Temperature, mean (SD), °C | 36.7 (0.5) | 36.7 (0.4) |

*Sampled into shifts = unique patients appearing in any of the 1,000 simulated shifts (11,630 of 12,020 test patients, 96.8%). Distributions are nearly identical, as expected from uniform random sampling without replacement within shifts. 1,000 shifts generated: 333 low-census (25 patients), 333 medium-census (40 patients), 334 high-census (60 patients), yielding 41,685 total patient-encounters. Abbreviations: BP, blood pressure; EMS, emergency medical services; ESI, Emergency Severity Index; SD, standard deviation;* $SpO_2$*, peripheral oxygen saturation.*

**Supplementary Table 3. Queue-ordering policies evaluated in this study.**

| Policy | Family | Pairwise judge | Capsule format | Site-specific training data |
|---|---|---|---|---|
| FIFO | Standard-of-care | – | – | Not required |
| ESI | Standard-of-care | – | – | Not required |
| NEWS2 | Standard-of-care | – | – | Not required |
| XGBoost | Supervised ML | – | Numeric features | 81,588 labeled encounters (Site A) |
| BT-Heuristic | BT pairwise | NEWS2 score | – | Not required |
| BT-LLM | BT pairwise | GPT-4.1 | Structured (~200 tokens) | Not required |
| BT-LLM-Enriched | BT pairwise | GPT-4.1 | Enriched (uncapped) | Not required |

*Abbreviations: BT, Bradley-Terry; ESI, Emergency Severity Index; FIFO, first-in-first-out; LLM, large language model; ML, machine learning; NEWS2, National Early Warning Score 2.*

**Supplementary Table 4. ESI-based top-5 identification rates for deteriorators by subgroup.**

| Subgroup | N deteriorator-obs. | ESI top-5 rate | NEWS2 top-5 rate | p-value (ESI) |
|---|---|---|---|---|
| **Age** | | | | <0.001 |
| < 40 years | 177 | 62.7% | 61.6% | |
| 40–64 years | 323 | 43.7% | 44.9% | |
| 65+ years | 622 | 43.6% | 44.5% | |
| **Sex** | | | | 0.89 |
| Female | 484 | 46.3% | 41.7% | |
| Male | 638 | 46.9% | 51.6% | |
| **Arrival mode** | | | | <0.001 |
| EMS | 674 | 53.4% | 51.5% | |
| Self | 403 | 31.8% | 40.2% | |

*Top-5 identification rate = proportion of deteriorators ranked in positions 1–5 under each policy. P-values from chi-squared tests for variation across subgroups within the ESI policy. N deteriorator-observations reflects patient-shift pairs (patients may appear in multiple shifts). Abbreviations: EMS, emergency medical services; ESI, Emergency Severity Index; NEWS2, National Early Warning Score 2.*

**Supplementary Table 5. Deterioration base rates by subgroup.**

| Subgroup | N patients | Deterioration events | Rate |
|---|---|---|---|
| **Age** | | | |
| < 40 years | 3,839 | 49 | 1.3% |
| 40–64 years | 4,219 | 90 | 2.1% |
| 65+ years | 3,962 | 162 | 4.1% |
| **Sex** | | | |
| Female | 6,411 | 135 | 2.1% |
| Male | 5,602 | 166 | 3.0% |

*Deterioration = composite outcome (ICU admission within 6 hours, intubation, vasopressor, mechanical ventilation within 6 hours of arrival, or in-hospital death). Rates computed on the chronological test split (12,020 visits). Abbreviation: ICU, intensive care unit.*

**Supplementary Figure 1. Sensitivity of median TTP-Det to wait-time model specification (MIMIC-IV-ED).**

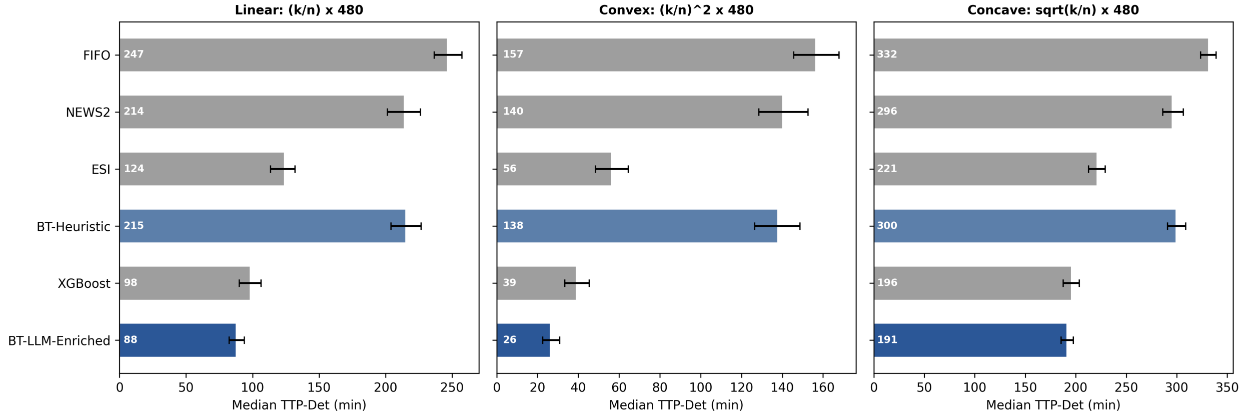


Figure 1: Median simulated time-to-provider for deteriorators recomputed under three monotonically increasing wait-time functions: linear ($(k/n) \times 480$ min, used in main analysis), convex ($(k/n)^2 \times 480$ min, modeling front-loaded throughput), and concave ($\sqrt{k/n} \times 480$ min, modeling back-loaded throughput). Per-patient queue positions from the MIMIC-IV-ED external validation (500 shifts) were held fixed; only the mapping from queue position to wait time was varied. The relative ordering of policies is preserved across all three models, confirming that between-policy comparisons are robust to the choice of wait-time function. Gray bars denote classification-based policies; blue bars denote context-aware pairwise ranking configurations. Error bars show 95% bootstrap confidence intervals. Abbreviations: BT, Bradley-Terry; ESI, Emergency Severity Index; FIFO, first-in-first-out; LLM, large language model; MIMIC-IV-ED, Medical Information Mart for Intensive Care IV Emergency Department; NEWS2, National Early Warning Score 2; TTP-Det, simulated time-to-provider for deteriorators.

**Supplementary Figure 2. Median and 95th-percentile simulated time-to-provider (TTP) for deteriorators across development (MC-MED, blue) and external validation (MIMIC-IV-ED, orange) datasets.**

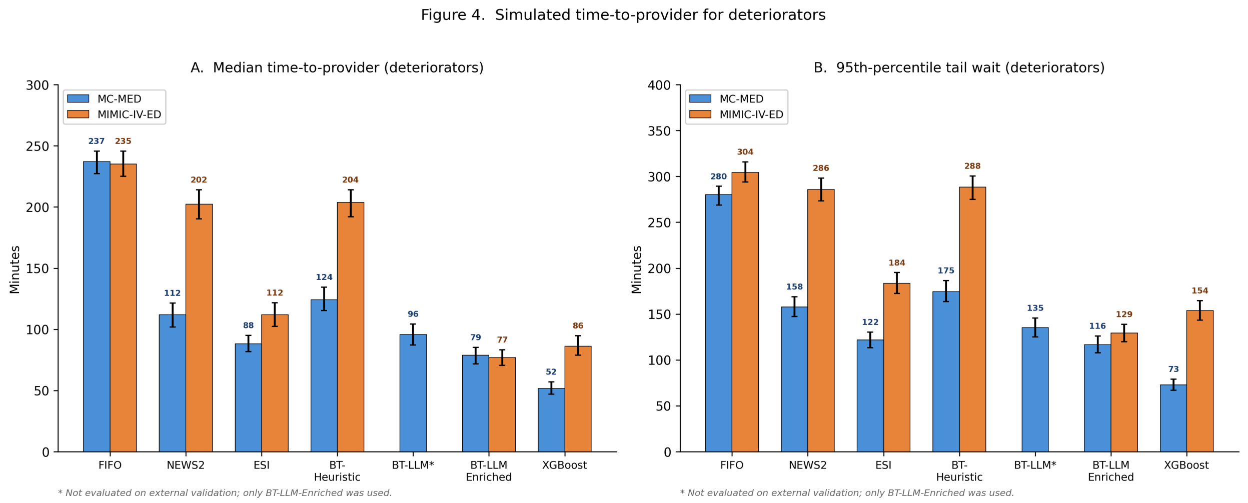


Figure 2: Lower values indicate earlier provider contact. Error bars show 95% bootstrap confidence intervals. Asterisk (*) indicates BT-LLM was not evaluated on external validation; only BT-LLM-Enriched was used. Abbreviations: BT, Bradley-Terry; ESI, Emergency Severity Index; FIFO, first-in-first-out; LLM, large language model; MC-MED, Multimodal Clinical Monitoring in the Emergency Department; MIMIC-IV-ED, Medical Information Mart for Intensive Care IV Emergency Department; NEWS2, National Early Warning Score 2; TTP, time-to-provider.

**Supplementary Table 6. Sensitivity of policy performance to deterioration outcome window (6, 12, and 24 hours).**

**Panel A: MC-MED (1,000 simulated shifts, Site A)**

| Policy | Window | Det. rate | Recall@5 | AUROC | Median TTP-Det (min) |
|---|---|---|---|---|---|
| FIFO | 6h | 2.5% | 0.119 | 0.493 | 237 |
| FIFO | 12h | 2.7% | 0.126 | 0.493 | 237 |
| FIFO | 24h | 2.7% | 0.126 | 0.494 | 237 |
| ESI | 6h | 2.5% | 0.491 | 0.811 | 88 |
| ESI | 12h | 2.7% | 0.477 | 0.799 | 94 |
| ESI | 24h | 2.7% | 0.476 | 0.798 | 94 |
| NEWS2 | 6h | 2.5% | 0.487 | 0.754 | 112 |
| NEWS2 | 12h | 2.7% | 0.484 | 0.753 | 113 |
| NEWS2 | 24h | 2.7% | 0.482 | 0.754 | 113 |
| XGBoost | 6h | 2.5% | 0.639 | 0.887 | 54 |
| XGBoost | 12h | 2.7% | 0.620 | 0.878 | 58 |
| XGBoost | 24h | 2.7% | 0.621 | 0.877 | 59 |
| BT-Heuristic | 6h | 2.5% | 0.400 | 0.710 | 133 |
| BT-Heuristic | 12h | 2.7% | 0.402 | 0.712 | 132 |
| BT-Heuristic | 24h | 2.7% | 0.402 | 0.713 | 132 |

**Panel B: MIMIC-IV-ED (500 simulated shifts, Site B)**

| Policy | Window | Det. rate | Recall@5 | AUROC | Median TTP-Det (min) |
|---|---|---|---|---|---|
| FIFO | 6h | 4.6% | 0.116 | 0.471 | 245 |
| FIFO | 12h | 6.2% | 0.123 | 0.476 | 243 |
| FIFO | 24h | 6.7% | 0.123 | 0.480 | 240 |
| ESI | 6h | 4.6% | 0.420 | 0.756 | 110 |
| ESI | 12h | 6.2% | 0.409 | 0.754 | 111 |
| ESI | 24h | 6.7% | 0.396 | 0.751 | 112 |
| NEWS2 | 6h | 4.6% | 0.285 | 0.568 | 202 |
| NEWS2 | 12h | 6.2% | 0.308 | 0.583 | 191 |
| NEWS2 | 24h | 6.7% | 0.309 | 0.585 | 189 |
| XGBoost | 6h | 4.6% | 0.499 | 0.817 | 83 |
| XGBoost | 12h | 6.2% | 0.510 | 0.817 | 82 |
| XGBoost | 24h | 6.7% | 0.508 | 0.817 | 81 |
| BT-Heuristic | 6h | 4.6% | 0.219 | 0.560 | 207 |

| Policy | Window | Det. rate | Recall@5 | AUROC | Median TTP-Det (min) |
|---|---|---|---|---|---|
| BT-Heuristic | 12h | 6.2% | 0.254 | 0.579 | 194 |
| BT-Heuristic | 24h | 6.7% | 0.252 | 0.580 | 192 |

*The relative ordering of policies was preserved across all three outcome windows at both sites. BT-LLM-Enriched was not re-evaluated because its rankings are derived from triage data via pairwise comparison and are independent of the outcome definition; the same queue orderings would apply under any outcome window. Abbreviations: AUROC, area under the receiver operating characteristic curve; BT, Bradley–Terry; Det., deterioration; ESI, Emergency Severity Index; FIFO, first-in-first-out; NEWS2, National Early Warning Score 2; TTP-Det, simulated time-to-provider for deteriorators.*

**Supplementary Material: LLM Prompt Template**

The following prompt template (version pairwise_v002) was used for all LLM-based pairwise comparisons (LLM, large language model; ED, emergency department). Temperature was set to 0. The `{capsule_A}` and `{capsule_B}` placeholders were replaced with the serialized triage capsule text for each patient pair.

**System message:**

> You are an emergency department triage prioritization system. You will be shown two patients (A and B) with their triage capsules. Your task: determine which patient should be seen by a provider FIRST based on near-term risk of clinical deterioration.
>
> OUTPUT FORMAT: Return ONLY a single JSON object with exactly these keys:
> winner_urgency: "A" | "B" | "TIE"
> p_win_bucket: 0.55 | 0.65 | 0.75 | 0.85
> confidence: "LOW" | "MED" | "HIGH"
> urgency_bucket_A: "0-15" | "15-60" | "60-180" | ">180"
> urgency_bucket_B: "0-15" | "15-60" | "60-180" | ">180"
> vertical_A: "YES" | "NO" | "UNCERTAIN"
> vertical_B: "YES" | "NO" | "UNCERTAIN"
> care_area_A: "RESUS" | "MONITORED" | "BED" | "VERTICAL" | "PSYCH_SAFE" | "ISOLATION" | "UNCERTAIN"
> care_area_B: "RESUS" | "MONITORED" | "BED" | "VERTICAL" | "PSYCH_SAFE" | "ISOLATION" | "UNCERTAIN"
> rationale_tags: [list of strings from ALLOWED TAGS ONLY]
>
> ALLOWED RATIONALE TAGS (use ONLY these):
> HYPOXIA, HYPOTENSION, SEPSIS_CONCERN, CHEST_PAIN_HIGH_RISK, NEURO_DEFICIT,
> BLEEDING, ALTERED_MENTAL_STATUS, FRAILTY_RISK, ANTICOAGULANT, SEVERE_PAIN_RED_FLAGS,
> MISSING_VITALS, STABLE, LOW_ACUITY, OTHER
>
> RULES:
> - Base your judgment ONLY on the information provided in each capsule.
> - Missing vitals (shown as UNKNOWN) should increase uncertainty and generally bias toward a more cautious (higher urgency) assessment.
> - Use TIE when the patients are comparably urgent.
> - p_win_bucket reflects how much more urgent the winner is: 0.55 = barely, 0.85 = substantially.
> - vertical = YES means the patient can be evaluated standing/sitting without a bed.
> - Do NOT output any text outside the JSON object.
>
> EXAMPLE OUTPUT:
> {"winner_urgency": "A", "p_win_bucket": 0.75, "confidence": "HIGH", "urgency_bucket_A": "0-15", "urgency_bucket_B": "60-180", "vertical_A": "NO", "vertical_B": "YES", "care_area_A": "MONITORED", "care_area_B": "VERTICAL", "rationale_tags": ["HYPOTENSION", "SEPSIS_CONCERN"]}

**User message template:**

Compare these two ED patients. Return a single JSON object.
PATIENT A:
{capsule_A}
PATIENT B:
{capsule_B}

### Appendix A. Bradley-Terry Update Dynamics

**Likelihood formulation.** For patients $A$ and $B$ with latent severity parameters $\theta_A$ and $\theta_B$, the Bradley-Terry model defines the probability that $A$ should be seen before $B$ as:

$$P(A \succ B) = \sigma(\theta_A - \theta_B) = \frac{1}{1 + e^{-(\theta_A - \theta_B)}}$$

where $\sigma(\cdot)$ is the logistic sigmoid function.

**Update equations.** After each pairwise comparison yielding outcome $y \in \{0, 0.5, 1\}$ (where $y = 1$ if $A$ wins, $y = 0$ if $B$ wins, and $y = 0.5$ for a tie), parameters are updated via stochastic gradient descent on the negative log-likelihood:

$$\theta_A \leftarrow \theta_A + \eta \cdot w \cdot (y - \hat{p})$$

$$\theta_B \leftarrow \theta_B - \eta \cdot w \cdot (y - \hat{p})$$

where $\hat{p} = \sigma(\theta_A - \theta_B)$ is the predicted probability, $\eta = 0.4$ is the learning rate, and $w$ is a confidence weight derived from the judge's self-reported confidence level:

$$w = \begin{cases} 0.4 & \text{if confidence} = \texttt{LOW} \\ 0.7 & \text{if confidence} = \texttt{MED} \\ 1.0 & \text{if confidence} = \texttt{HIGH} \end{cases}$$

The confidence weighting downweights updates from uncertain comparisons, reducing the influence of ambiguous pairwise judgments on the ranking.

**Initialization and reset.** All $\theta$ values are initialized to 0 at the start of each simulated shift. There is no carry-over of learned parameters between shifts; each shift represents an independent ranking episode. In a real-time deployment, $\theta$ values would similarly reset at shift boundaries or when the waiting room clears.

**Regularization.** No explicit regularization term was applied. $\theta$ values are implicitly bounded by the small number of comparisons per patient (3–7 under the sentinel strategy). With $\eta = 0.4$, $w \leq 1.0$, and gradient magnitude $|y - \hat{p}| \leq 1.0$, a single comparison can shift $\theta$ by at most 0.4. Over 7 comparisons, the maximum $|\theta|$ is approximately 2.8, preventing unbounded growth.

**Sentinel selection strategy.** When a new patient arrives, sentinels are selected at evenly spaced quantiles of the current $\theta$ distribution among waiting patients. The new patient is first compared against the median sentinel, then against additional sentinels (up to 4 total comparisons). If the new patient's $\theta$, after sentinel comparisons, places them in the top 5, two additional comparisons against other top-ranked patients are performed (top-$k$ reinforcement). This yields 3–7 comparisons per arrival, rather than the $O(n)$ comparisons required by exhaustive pairwise comparison.

**Stability under non-transitive judgments.** The Bradley-Terry model produces a consistent global ranking even when individual pairwise comparisons are non-transitive, which is a known advantage of this framework over sorting-based approaches that assume transitivity. Non-transitive cycles ($A \succ B \succ C \succ A$) do not cause divergence; instead, the maximum likelihood estimates assign

$\theta$ values that best explain the observed comparison outcomes in aggregate. In the online SGD setting used here, non-transitive comparisons may introduce oscillation in individual $\theta$ values, but the ranking order (determined by the relative magnitudes of all $\theta$ values) remains stable because each patient's estimate is informed by multiple independent comparisons against different sentinels.

**Learning rate sensitivity.** Hyperparameter sensitivity was evaluated using the heuristic (NEWS2) judge on the development dataset (MC-MED, 1,000 shifts). Because the BT update mechanism is independent of the comparison function, these results characterize the ranking algorithm's sensitivity to its own parameters rather than the judge's. The learning rate was varied over $\{0.1, 0.2, 0.3, 0.4, 0.5, 0.6, 0.8\}$. Recall@5 ranged from 0.411 to 0.425 (spread: 1.4 percentage points); AUROC ranged from 0.704 to 0.709 (spread: 0.5 points). Performance was essentially flat across the full range, indicating that the BT ranking is not sensitive to learning rate selection. The value $\eta = 0.4$ was adopted as a standard midpoint.

| Learning rate ($\eta$) | Recall@5 | AUROC | Median TTP-Det (min) |
|---|---|---|---|
| 0.1 | 0.414 | 0.704 | 135.4 |
| 0.2 | 0.421 | 0.705 | 135.8 |
| 0.3 | 0.425 | 0.709 | 133.6 |
| 0.4 | 0.411 | 0.706 | 134.7 |
| 0.5 | 0.412 | 0.706 | 134.8 |
| 0.6 | 0.419 | 0.706 | 134.6 |
| 0.8 | 0.416 | 0.709 | 133.3 |

**Sentinel count sensitivity.** The number of sentinels was varied over $\{3, 4, 5, 6, 7\}$ with $\eta = 0.4$. AUROC was stable (0.700–0.708, spread: 0.8 points). Recall@5 showed modest variation (0.397–0.444), reflecting stochastic differences in which patients are selected as sentinels at smaller counts, but the overall ranking quality (AUROC, TTP) was robust. The value 5 was adopted as a balance between comparison cost and ranking resolution.

| Sentinel count | Recall@5 | AUROC | Median TTP-Det (min) |
|---|---|---|---|
| 3 | 0.397 | 0.700 | 138.0 |
| 4 | 0.430 | 0.705 | 135.9 |
| 5 | 0.411 | 0.706 | 134.7 |
| 6 | 0.444 | 0.708 | 134.9 |
| 7 | 0.412 | 0.707 | 135.2 |

*Abbreviations used in this appendix: AUROC, area under the receiver operating characteristic curve; BT, Bradley-Terry; MC-MED, Multimodal Clinical Monitoring in the Emergency Department; NEWS2, National Early Warning Score 2; SGD, stochastic gradient descent; TTP-Det, simulated time-to-provider for deteriorators.*
